# Gut microbiota signatures differentiate trajectory-defined response phenotypes and predict self-management outcomes in irritable bowel syndrome

**DOI:** 10.64898/2026.05.18.26353470

**Authors:** Jie Chen, Aolan Li, Weizi Wu, Wanli Xu, Tingting Zhao, Angela R. Starkweather, Leonel Rodriguez, Ming-Hui Chen, Xiaomei S. Cong

**Affiliations:** College of Nursing, Florida State University, Tallahassee, FL 32306, USA; Yale School of Nursing, Orange, CT 06477, USA; Department of Statistics, University of Connecticut, Storrs, CT 06269, USA; School of Nursing, University of Connecticut, Storrs, CT 06269, USA; School of Nursing, Columbia University, New York, NY 10032, USA; Division of Nursing Science, Rutgers School of Nursing, New Brunswick, NJ 08901, USA; Yale School of Medicine, New Haven, CT 06510, USA

**Keywords:** irritable bowel syndrome, chronic pain, machine learning, precision health, gut microbiota

## Abstract

**Background:** Heterogeneity in symptom presentation and treatment response in irritable bowel syndrome (IBS) remains poorly understood. The gut microbiota may contribute to this variability, but its role in shaping symptom trajectories and responses to self-management interventions is unclear.

**Objective:** To identify symptom trajectory phenotypes and determine whether gut microbiota composition and function distinguish these phenotypes and predict multidimensional responses to pain self-management interventions in young adults with IBS.

**Design:** Ancillary data analysis from a randomized control trial (NCT03332537).

**Methods:** Participants with longitudinal data (n = 62) were analyzed using longitudinal k-means clustering (KML) based on trajectories of measures in IBS quality of life (QOL), Brief Pain Inventory (BPI), and psychoneurological outcomes (anxiety, applied cognition, depression, fatigue, global health, positive affect, and sleep disturbance) over 12 weeks. Baseline differences between clusters were assessed with Wilcoxon rank-sum tests, and longitudinal changes were evaluated with linear mixed models. Gut microbiota composition and predicted functional pathways were compared between phenotypes. Bayesian Additive Regression Trees (BART) models were used to identify baseline microbial taxa and pathways predictive of longitudinal changes in QOL, BPI pain interference, and severity.

**Results:** Two distinct trajectory-defined response phenotypes were identified: a Constrained Response Phenotype (Phenotype A, n = 35) and an Adaptive Multidomain Response Phenotype (Phenotype B, n = 27). At baseline, Phenotype B showed lower pain severity and interference, but higher levels of anxiety, depression, and fatigue compared to Phenotype A. Over 12 weeks, both phenotypes showed improvements in pain outcomes (all p < 0.05), but only Phenotype B demonstrated broad improvements across psychoneurological domains and QOL (all p < 0.05). Phenotype A exhibited more limited improvements and worsening in several psychoneurological domains. Gut microbiota functional pathways differed between phenotypes, including pathways related to xenobiotic degradation, amino acid metabolism, bile secretion, and immune-related processes (all raw p < 0.05), although these did not remain significant after multiple testing correction. Machine learning models identified distinct, phenotype-specific microbial predictors of intervention response. In Phenotype A, genera such as *Alistipes* and *Sutterella* were consistently identified across models, whereas in Phenotype B, predictors included *Phascolarctobacterium*, *Collinsella*, and *Parabacteroides*. Functional pathways also differed between phenotypes, suggesting distinct microbiome-linked mechanisms underlying symptom trajectories and responses to pain interventions.

**Conclusions:** Young adults with IBS exhibit distinct multidimensional response phenotypes that are associated with differential clinical and microbiome profiles. Baseline gut microbiota composition and functional capacity demonstrate phenotype-specific predictive signatures of treatment response, supporting a microbiome-informed framework for stratifying patients and advancing personalized self-management strategies in IBS.

**WHAT IS KNOWN:**

□ Substantial heterogeneity exists in irritable bowel syndrome symptoms and treatment response, with variability across pain, psychological distress, and quality of life domains.
□ Gut microbiota composition and function are linked to IBS pathophysiology, including associations with pain sensitivity, inflammation, and brain–gut signaling.
□ Self-management interventions (e.g., mindfulness, behavioral strategies) can improve IBS symptoms, but responses are inconsistent and difficult to predict.

**WHAT IS NEW HERE:**

□ Distinct longitudinal symptom trajectory phenotypes were identified, separating individuals with pain-predominant and limited psychoneurological improvement from those with lower pain and greater multidomain psychoneurological improvement.
□ Gut microbiota composition and functional profiles varied between trajectory-defined clusters, indicating a biological foundation for differences in symptom patterns.
□ Machine learning models showed that gut microbiota features predict pain severity, interference, quality of life, and their longitudinal changes, supporting microbiome-based stratification for self-management outcomes.

## 1. Introduction

Irritable bowel syndrome (IBS) is a prevalent disorder of gut–brain interaction affecting up to 20% of adults worldwide,^1–5^ typically manifests in early adulthood and affects more women than men.^1–7^ It is a leading cause of healthcare utilization and work absenteeism, accounting for more than $21 billion annually in direct and indirect costs in the United States alone.^8–12^ Despite its high prevalence and economic burden, effective long-term management remains limited.^13–15^ Pharmacologic therapies yield modest and inconsistent benefits and often fail to address the multidimensional nature of IBS, including co-occurring pain, psychological distress, and impaired quality of life (QOL).^16–20^ Patients with IBS frequently report prolonged difficulty identifying effective self-management strategies, contributing to sustained pain and reduced QOL.^21–24^ These challenges are further compounded by substantial inter-individual variability in symptom presentation and treatment response, highlighting the need to identify distinct response patterns that may inform more precise, mechanism-based, non-pharmacologic approaches within a whole-person health framework.

The gut microbiome represents a central pathway linking self-management behaviors to symptom regulation in IBS.^45,75–77^ Beyond taxonomic composition, microbial functional capacity, such as pathways involved in short-chain fatty acid production, aromatic amino acid metabolism, bile acid transformation, and neuroactive compound synthesis, plays a critical role in modulating host neuroimmune and neuroendocrine processes.^78–81^ These microbial metabolites influence vagal signaling, inflammation, intestinal permeability, and gastrointestinal motility, all of which are implicated in IBS pathophysiology. In parallel, stress-related alterations in autonomic and immune function can reshape the gut environment and microbial activity, suggesting a bidirectional regulatory system. Self-management interventions, particularly those targeting stress and behavioral regulation, may act through this gut–brain axis by inducing downstream shifts in microbial function. However, whether variability in microbial composition and functional capacity contributes to differences in longitudinal symptom trajectories and intervention responsiveness remains poorly understood.

A critical gap remains in understanding how heterogeneity in multidimensional symptom trajectories relates to underlying biological mechanisms, particularly microbial functional dynamics. IBS is characterized by substantial inter-individual variability across pain, psychological symptoms, and quality of life, yet most prior studies rely on average treatment effects and do not capture distinct patterns of response over time. Person-centered approaches, such as trajectory-based clustering, offer an opportunity to identify subgroups of individuals with distinct longitudinal response profiles that integrate these domains. Identification of such phenotypes may provide a more clinically meaningful representation of intervention response and uncover latent regulatory differences not apparent in group-level analyses. Moreover, the extent to which gut microbiota composition and function distinguish these trajectory-defined phenotypes, and whether such differences reflect underlying regulatory mechanisms rather than treatment assignment, remains unclear.

To address these gaps, this study leveraged longitudinal data from a randomized controlled trial to: (1) identify multidimensional symptom trajectory phenotypes using repeated measures of QOL, pain, and psychoneurological outcomes; (2) determine whether gut microbiota composition and inferred functional pathways differ between these phenotypes; and (3) evaluate whether baseline microbial features predict intervention response using a machine learning framework. By integrating longitudinal phenotyping, microbiome profiling, and predictive modeling, this study aims to advance microbiome-informed stratification approaches for personalized self-management in IBS and to characterize response heterogeneity beyond treatment-group effects.

## 2. Methods

### 2.1 Study Design and Participants

Details of the design, rationale, and primary results of the RCT (NCT03332537) have been published.^25,26^ The RCT protocol was approved by the University Institute Review Board (IRB). Informed consent was acquired from each participant. Privacy was maintained during the entire data collection and management process. Young adults with IBS (N = 80) were recruited in the northeastern United States for this RCT. Young adults were enrolled in the study if they were (1)18-29 years of age; (2) able to read and speak in English; (3) able to access the internet; (4) having a clinical diagnosis of IBS according to the Rome III or IV criteria from a healthcare provider (for IBS subjects only). Subjects were excluded if they had (1) chronic pain conditions (e.g., chronic pelvic pain, headache, back pain, etc.); (2) severe mental health conditions; (3) celiac disease or inflammatory bowel disease; (4) infectious diseases; (5) diabetes mellitus; (6) injury or open skin lesions on the non-dominant arm; (7) history of substance abuse or regular opioid use; (8) history of prebiotics/probiotics or antibiotics use in the past months.^26^ Women during pregnancy or within 3 months postpartum period were also excluded. Eligible participants with IBS were allocated to the intervention groups (Online Modules Plus vs. Online Modules) using a stratified, blocked randomization scheme. The full description of randomization and blinding was previously described in the published study protocol.^25^ De-identified data were used in this analysis.

### 2.2 Assessment of Variables

This analysis leveraged data collected in the RCT (NCT03332537).^25,26^ Measurements in the RCT (NCT03332537) included demographic characteristics, pain, quality of life, and psychoneurological symptoms. The demographic characteristics include age, sex, ethnicity, race, education level, and other factors measured using the National Institute of Nursing Research common data elements.^27^ Questionnaires were completed online through the REDCap system using a laptop or iPad. Participants with IBS completed all questionnaires at enrollment (W0, 0 weeks) and at 6- and 12-week follow-up visits (W6 and W12, respectively).

#### Pain

Pain severity and interference were measured using the Brief Pain Inventory (BPI). The BPI uses 0-10 rating scales, with higher scores indicating more severe pain.^28^

#### Quality of life

IBS quality of life was measured by a 34-item IBS-specific quality of life instrument.^29^ This instrument was designed to capture patients’ perceptions of their daily functions interfered with by IBS. There are 8 subscales in this five-point Likert instrument. The score range for this scale is 0-100, with higher scores indicating better quality of life.

#### IBS-related psychoneurological measures

IBS-related psychoneurological outcomes, including anxiety, applied cognition, depression, fatigue, global health, positive affect, and sleep disturbance, were measured using the NIH Patient-Reported Outcomes Measurement Information System (PROMIS®) and scored according to the system instructions.^30^ A higher T score on the PROMIS measurement indicated a higher intensity of the measured concept, and a mean score greater than 55 indicates that a study subject experienced a significantly higher intensity of the symptom/well-being than the healthy reference population, according to the PROMIS guide.

#### Stool sample collection and gut microbiota sequencing

Participants collected stool samples at home following our previous protocol using the OMNIgene®-gut (DNA Genotek, Ottawa, Canada) kit.^31,32^ Training on collecting the samples at home was provided to the participants by research personnel. After collection, participants were instructed to mail the samples back to the research laboratory within a week of the baseline visit, using a pre-labeled, pre-paid envelope with a tracking barcode. Once received by the laboratory, the stool samples were aliquoted into small tubes and stored at -80 ℃ in a freezer for bulk processing. A total of 0.25 g of stool from each participant was placed into a bead tube and sent out for DNA extraction and sequencing. The 16S rRNA V4 region amplicon was sequenced by the Illumina MiSeq 2000 platform (Illumina, San Diego, CA) at the University of Connecticut (UConn) Microbial Analysis, Resource, and Services laboratory following our previous protocol.^32^ The raw 16S rRNA sequencing data were processed by the Mothur 1.43.0 software following the analysis pipeline of Miseq (http://www.mothur.org/wiki/MiSeq_SOP) to obtain the taxonomy and diversity of the gut microbiota. Paired-end sequences were combined into contigs and aligned against the SILVA 132 V4 16S rRNA gene reference alignment database, with poor-quality sequences removed. Operational taxonomic units (OTUs) were identified at a 97% identity.^33,34^

Microbial functional profiles were inferred from 16S rRNA gene sequences using Tax4Fun2, which predicts gene family abundances based on reference genomes.^35^ Predicted functions were annotated as Kyoto Encyclopedia of Genes and Genomes orthologs (KOs) and aggregated for downstream pathway-level analyses.^36^

### 2.3 Data analysis

All statistical analyses were conducted using R version 4.5.1. Longitudinal k-means clustering (KML)^37,38^ was applied to the IBS quality of life (QOL), Brief Pain Inventory (BPI), and psychoneurological outcomes (anxiety, applied cognition, depression, fatigue, global health, positive affect, and sleep disturbance) measured by the Patient-Reported Outcomes Measurement Information System (PROMIS) over 12 weeks.

Exploratory analyses in longitudinal gut microbiota features, including the diversity indices, abundance of genus, and predicted pathways among participants with available stool samples at weeks 0, 6, and 12, were examined using limma.^39^ The interaction term (cluster × visit) of gut microbiota features was also tested by limma models.^39^ Exploratory genus-level differential abundance analyses using ANCOM-BC2.^40^ Pathways highlighted by phenotype and visit interaction were visualized in a heatmap.

Bayesian Additive Regression Trees (BART)^41^ were employed to investigate the relationship between predictors and the outcomes of QOL, BPI pain interference, and pain severity. A forward prediction model estimates the change in outcomes from baseline to a future visit using gut microbiota predictors from baseline visit (e.g., baseline predictors were used to predict the 6-week and 12-week changes from baseline). The predictor variables included gut microbiota composition (feature abundances) and predicted functional profiles (KOs pathways). For the outcomes, positive values indicated an increase from the earlier to the later visit, whereas negative values indicated a decrease. The models did not explicitly model within-subject correlation across visits. Longitudinal information was incorporated only through the continuous change scores. Within each subgroup, we applied an unsupervised prevalence filter, retaining features present in more than 10% of samples. Presence was defined as abundance > 0 for genus-level taxa and relative abundance > 5 × 10^-3^ for predicted KEGG level 3 pathways. KEGG features were then CLR-transformed with a small pseudocount as needed. We applied 3-fold cross-validation across various BART hyperparameter settings (number of trees, node prior, and tree structure priors), selecting the configuration with the lowest mean out-of-sample RMSE. This configuration was then refit to the full dataset for each subgroup and follow-up time point. Variable importance was summarized by variable inclusion proportions, and model fit was described using in-sample RMSE and RMSE reduction relative to an intercept-only model. The top 10 influential predictors were identified based on their variable inclusion proportions, defined as the proportion of times a variable was used to split a decision node across all trees. The MCMC process included a burn-in period of 10,000 iterations, followed by posterior sampling with every 20th draw retained (thinning), yielding 10,000 posterior samples. Convergence was evaluated using trace plots and autocorrelation functions, confirming adequate mixing and stability of posterior draws.

## 3. Results

### 3.1 Longitudinal multidimensional trajectories-based phenotypes of self-management outcomes in young adults with IBS

Among the 80 participants enrolled in the RCT, 18 contributed only baseline data and could not be assigned to a trajectory cluster (unclassified, baseline-only; n = 18). Of the participants with longitudinal data, 62, who had at least one follow-up, were classified into two multidimensional response trajectories using KML: “Constrained Response Phenotype (Phenotype A, n = 35)” and “Adaptive Multidomain Response Phenotype (Phenotype B, n = 27).” Table 1 presents the demographic characteristics of participants in Phenotype A and B and those unclassified at baseline. There was no significant difference in demographic characteristics between the two phenotypes and those unclassified at baseline.

**Table 1.** Baseline demographic characteristics.

|  | Phenotype A (n = 35) | Phenotype B (n = 27) | Unclassified (n = 18) | p-value |
| --- | --- | --- | --- | --- |
| Age, Mean (SD) | 21.83 (2.98) | 21.56 (2.38) | 20.28 (1.64) | 0.105 |
| Year of IBS diagnosis, Mean (SD) | 3.49 (3.63) | 2.59 (2.21) | 3.17 (2.79) | 0.517 |
| Sex, n (%) |  |  |  | 0.644 |
| Female | 25 (71.43%) | 22 (81.48%) | 14 (77.78%) |  |
| Male | 10 (28.57%) | 5 (18.52%) | 4 (22.22%) |  |
| Race, n (%) |  |  |  | 0.195 |
| Asian | 5 (14.29%) | 4 (14.81%) | 1 (5.56%) |  |
| Black or African American | 6 (17.14%) | 0 (0.0%) | 2 (11.11%) |  |
| White | 24 (68.57%) | 23 (85.19%) | 15 (83.33%) |  |
| Ethnicity, n (%) |  |  |  | 0.466 |
| Hispanic or Latino | 3 (8.57%) | 3 (11.11%) | 1 (5.56%) |  |
| Not Hispanic or Latino | 28 (80.00%) | 23 (85.19%) | 17 (94.44%) |  |
| Not reported | 4 (11.43%) | 1 (3.70%) | 0 (0.0%) |  |
| Education, n (%) |  |  |  |  |
| High school or below | 4 (11.43%) | 3 (11.12%) | 0 |  |
| College or associate degree | 18 (51.42%) | 12 (44.44%) | 16 (88.89%) |  |
| Bachelor's degree | 5 (14.29%) | 6 (22.22%) | 2 (11.11%) |  |
| Graduate or Doctorate degree | 8 (22.86%) | 6 (22.22%) | 0 |  |
| Primary Caregiver, n (%) |  |  |  |  |
| Self-care | 16 (45.71%) | 16 (59.26%) | 8 (44.44%) |  |
| Cared by others | 19 (54.29%) | 11 (40.74%) | 10 (55.56%) |  |
| Employment Status, n (%) |  |  |  | 0.159 |
| Student | 23 (65.71%) | 23 (85.19%) | 11 (61.11%) |  |
| Working now | 11 (31.43%) | 4 (14.81%) | 5 (27.78%) |  |
| Unemployed or other | 1 (2.86%) | 0 (0.0%) | 2 (11.11%) |  |
| Marital Status, n (%) |  |  |  | 0.584 |
| Never married | 2 (5.71%) | 1 (3.70%) | 0 (0.0%) |  |
| Married | 33 (94.29%) | 26 (96.30%) | 18 (100.00%) |  |
| Family member with IBS, n (%) |  |  |  | 0.966 |
| No | 24 (68.57%) | 19 (70.37%) | 12 (66.67%) |  |
| Yes | 11 (31.43%) | 8 (29.63%) | 6 (33.33%) |  |
| Intervention groups, n (%) |  |  |  | 0.057 |
| Online Module | 23 (65.71%) | 12 (44.44%) | 6 (33.33%) |  |
| Online Module plus Nurse-led Support | 12 (34.29%) | 15 (55.56%) | 12 (66.67%) |  |

Note: Phenotype A, Constrained Response Phenotype; Phenotype B, Adaptive Multidomain Response Phenotype; Unclassified, no follow-up data available.
<sup>1</sup>P-values are to compare the demographic variables between Phenotype A and Phenotype B using the Wilcoxon rank-sum test for the numerical variables and the Fisher's exact test for the categorical variables.

Phenotype A exhibits higher pain levels with limited psychoneurological improvement, while Phenotype B shows lower pain with broad psychoneurological gains. Using Wilcoxon rank-sum tests, Phenotype B had lower BPI interference and severity than Cluster A (p = 0.06 and p = 0.24, respectively), but higher baseline scores in anxiety, depression, and fatigue T-scores (all p < 0.05, Table 2). Longitudinal changes from baseline to 12 weeks were assessed with LMM (Table 3). Compared to baseline, both phenotypes improved in BPI outcomes at 12 weeks (Figure 1, all p < 0.05). Additionally, Phenotype B showed improvements across multiple domains, including better QOL, global health, and positive affect; and reductions in anxiety, applied cognition, depression, fatigue, and sleep disturbance (Figure 1, all p < 0.05). Conversely, Phenotype A exhibited more limited improvements, with several domains worsening (e.g., higher anxiety, applied cognition, depression, fatigue, and lower positive affect; all p < 0.05), while QOL saw a modest improvement (p = 0.07). Sleep disturbance showed distinct trajectories, decreasing in Phenotype B and increasing in Phenotype A (p = 0.053).

**Table 2.** Quality of life, pain, and psychoneurological comparison by phenotype across visits.

| Outcome | Week 0 |  |  |  | Week 0 |  |  |
| --- | --- | --- | --- | --- | --- | --- | --- |
|  | A (N=35) | B (N=27) | Not assigned (N = 18) | p | Not assigned (N = 18) | Phenotype A/B (N=62) | p |
| BPI interference | 2.61 ± 2.13 | 1.66 ± 1.58 | 2.12 ± 1.55 | 0.128 † | 2.12 ± 1.55 | 2.19 ± 1.95 | 0.725 † |
| BPI severity | 2.94 ± 1.69 | 2.38 ± 1.24 | 2.43 ± 0.99 | 0.440 † | 2.43 ± 0.99 | 2.69 ± 1.53 | 0.777 † |
| Sleep T-score | 50.78 ± 7.33 | 51.43 ± 7.99 | 51.46 ± 8.88 | 0.933 | 51.46 ± 8.88 | 51.06 ± 7.57 | 0.851 |
| Fatigue T-score | 52.12 ± 7.55 | 57.30 ± 8.07 | 58.24 ± 7.76 | 0.008 † | 58.24 ± 7.76 | 54.38 ± 8.14 | 0.077 |
| Cognition T-score | 34.32 ± 6.86 | 37.66 ± 7.04 | 36.59 ± 7.58 | 0.164 † | 36.59 ± 7.58 | 35.77 ± 7.08 | 0.665 † |
| Anxiety T-score | 57.30 ± 8.69 | 62.41 ± 7.73 | 61.51 ± 9.47 | 0.051 | 61.51 ± 9.47 | 59.53 ± 8.61 | 0.403 |
| Depression T-score | 49.33 ± 8.77 | 54.36 ± 7.97 | 50.32 ± 9.72 | 0.092 † | 50.32 ± 9.72 | 51.52 ± 8.73 | 0.668 † |
| Positive affect T-score | 53.95 ± 7.78 | 52.51 ± 5.59 | 53.88 ± 6.71 | 0.684 | 53.88 ± 6.71 | 53.32 ± 6.90 | 0.760 |
| Global health T-score | 47.86 ± 6.73 | 49.56 ± 5.87 | 47.57 ± 7.24 | 0.508 | 47.57 ± 7.24 | 48.60 ± 6.38 | 0.560 |

| Outcome | Week 0 |  |  | Week 6 |  |  | Week 12 |  |  |
| --- | --- | --- | --- | --- | --- | --- | --- | --- | --- |
|  | A (N=35) | B (N=27) | p | A (N=35) | B (N=27) | p | A (N=33) | B (N=23) | p |
| QOL-R | 65.90 ± 22.98 | 69.88 ± 17.47 | 0.701 † | 67.06 ± 22.52 | 78.51 ± 14.39 | 0.050 † | 69.05 ± 23.92 | 82.77 ± 13.06 | 0.029 † |
| BPI interference | 2.61 ± 2.13 | 1.66 ± 1.58 | 0.061 † | 2.36 ± 1.79 | 1.24 ± 1.50 | 0.007 † | 1.57 ± 1.84 | 0.81 ± 0.87 | 0.093 † |
| BPI severity | 2.94 ± 1.69 | 2.38 ± 1.24 | 0.156 | 2.45 ± 1.35 | 1.92 ± 1.26 | 0.099 † | 2.23 ± 1.55 | 1.49 ± 1.13 | 0.078 † |
| Sleep T-score | 50.78 ± 7.33 | 51.43 ± 7.99 | 0.741 | 53.49 ± 9.02 | 48.96 ± 7.67 | 0.041 | 52.69 ± 8.59 | 48.98 ± 8.10 | 0.033 † |
| Fatigue T-score | 52.12 ± 7.55 | 57.30 ± 8.07 | 0.007 † | 57.19 ± 8.83 | 53.86 ± 8.30 | 0.136 | 54.72 ± 8.92 | 51.13 ± 10.27 | 0.170 |
| Cognition T-score | 34.32 ± 6.86 | 37.66 ± 7.04 | 0.065 | 37.79 ± 7.77 | 35.20 ± 6.54 | 0.168 | 38.69 ± 7.92 | 33.77 ± 6.12 | 0.016 |
| Anxiety T-score | 57.30 ± 8.69 | 62.41 ± 7.73 | 0.019 | 61.25 ± 5.83 | 58.49 ± 7.22 | 0.101 | 59.96 ± 8.25 | 55.01 ± 8.08 | 0.030 |
| Depression T-score | 49.33 ± 8.77 | 54.36 ± 7.97 | 0.023 | 53.19 ± 7.98 | 49.85 ± 7.81 | 0.104 | 52.55 ± 8.70 | 47.82 ± 7.87 | 0.071 † |
| Positive affect T-score | 53.95 ± 7.78 | 52.51 ± 5.59 | 0.419 | 52.69 ± 7.96 | 55.19 ± 6.18 | 0.182 | 51.99 ± 7.67 | 55.82 ± 5.88 | 0.049 |
| Global health T-score | 47.86 ± 6.73 | 49.56 ± 5.87 | 0.301 | 49.02 ± 6.18 | 51.59 ± 6.59 | 0.120 | 49.09 ± 6.51 | 52.05 ± 5.77 | 0.086 |

Notes: Values are presented as mean ± SD. In the first block, baseline outcomes are compared across Phenotype A, Phenotype B, and participants with baseline data only who were not cluster-assigned. P-values compare Phenotype A and Phenotype B within each week using either a two-sample t-test or Wilcoxon rank-sum test, selected according to the Shapiro–Wilk normality assessment. For the three-group baseline comparison, one-way ANOVA or Kruskal–Wallis test was used. † nonparametric tests; otherwise parametric tests.

**Table 3.** Linear mixed-effects model results for quality of life, pain, and psychoneurological outcomes.

**Block A**
| Term | QOL_R |  |  | bpi_int |  |  | bpi_severity |  |  |
| --- | --- | --- | --- | --- | --- | --- | --- | --- | --- |
|  | Estimate | SE | p | Estimate | SE | p | Estimate | SE | p |
| Intercept | 51.018 | 23.571 | 0.034 | 5.407 | 1.811 | 0.004 | 5.899 | 1.425 | <0.001 |
| Phenotype B | 5.306 | 5.230 | 0.314 | -1.056 | 0.442 | 0.019 | -0.652 | 0.349 | 0.065 |
| Week 6 | 1.447 | 1.924 | 0.453 | -0.238 | 0.277 | 0.393 | -0.450 | 0.222 | 0.045 |
| Week 12 | 3.556 | 1.965 | 0.073 | -1.043 | 0.283 | <0.001 | -0.646 | 0.226 | 0.005 |
| Age | 1.211 | 0.939 | 0.202 | -0.141 | 0.071 | 0.052 | -0.081 | 0.056 | 0.155 |
| Years since IBS diagnosis | 0.040 | 0.647 | 0.951 | -0.079 | 0.055 | 0.157 | -0.146 | 0.044 | 0.001 |
| Female | -11.659 | 5.138 | 0.025 | 0.373 | 0.430 | 0.388 | -0.298 | 0.339 | 0.381 |
| White | -3.162 | 6.077 | 0.605 | 0.075 | 0.458 | 0.870 | 0.063 | 0.360 | 0.862 |
| Not Hispanic/Latino | -3.668 | 8.212 | 0.657 | 0.428 | 0.611 | 0.487 | -0.397 | 0.480 | 0.412 |
| Student | 3.294 | 2.840 | 0.248 | -0.229 | 0.335 | 0.496 | -0.240 | 0.266 | 0.368 |
| clusterB:factor(visit)6 | 7.369 | 2.915 | 0.013 | -0.159 | 0.420 | 0.705 | 0.032 | 0.336 | 0.925 |
| clusterB:factor(visit)12 | 7.971 | 3.057 | 0.010 | 0.281 | 0.439 | 0.523 | -0.141 | 0.351 | 0.688 |

**Block B**
| Term | Anxiety_TScore |  |  | Cognition_TScore |  |  | Depression_TScore |  |  |
| --- | --- | --- | --- | --- | --- | --- | --- | --- | --- |
|  | Estimate | SE | p | Estimate | SE | p | Estimate | SE | p |
| Intercept | 71.247 | 8.259 | <0.001 | 43.918 | 8.308 | <0.001 | 59.562 | 9.660 | <0.001 |
| Phenotype B | 5.555 | 1.963 | 0.006 | 3.948 | 1.905 | 0.041 | 5.078 | 2.170 | 0.022 |
| Week 6 | 3.830 | 1.133 | <0.001 | 3.478 | 0.937 | <0.001 | 3.816 | 0.925 | <0.001 |
| Week 12 | 2.350 | 1.156 | 0.044 | 4.114 | 0.956 | <0.001 | 3.057 | 0.945 | 0.002 |
| Age | -0.733 | 0.325 | 0.028 | -0.409 | 0.329 | 0.218 | -0.564 | 0.383 | 0.146 |
| Years since IBS diagnosis | 0.473 | 0.250 | 0.062 | 0.353 | 0.245 | 0.154 | 0.322 | 0.276 | 0.246 |
| Female | 1.858 | 1.946 | 0.343 | -1.845 | 1.918 | 0.339 | -0.143 | 2.176 | 0.948 |
| White | -0.617 | 2.094 | 0.769 | -2.249 | 2.119 | 0.293 | 0.914 | 2.476 | 0.713 |
| Not Hispanic/Latino | 0.523 | 2.801 | 0.853 | 0.439 | 2.846 | 0.878 | 0.634 | 3.337 | 0.850 |
| Student | -1.497 | 1.438 | 0.299 | 0.848 | 1.277 | 0.508 | -0.210 | 1.320 | 0.874 |
| clusterB:factor(visit)6 | -8.032 | 1.717 | <0.001 | -6.003 | 1.420 | <0.001 | -8.453 | 1.402 | <0.001 |
| clusterB:factor(visit)12 | -9.392 | 1.796 | <0.001 | -7.652 | 1.487 | <0.001 | -9.523 | 1.470 | <0.001 |

**Block C**
| Term | Fatigue_TScore |  |  | GlobHealth_TScore |  |  | PosiAffect_TScore |  |  |
| --- | --- | --- | --- | --- | --- | --- | --- | --- | --- |
|  | Estimate | SE | p | Estimate | SE | p | Estimate | SE | p |
| Intercept | 75.010 | 9.679 | <0.001 | 43.170 | 7.283 | <0.001 | 50.911 | 8.386 | <0.001 |
| Phenotype B | 5.403 | 2.237 | 0.018 | 1.596 | 1.675 | 0.343 | -1.586 | 1.867 | 0.398 |
| Week 6 | 5.108 | 1.149 | <0.001 | 1.133 | 0.837 | 0.179 | -1.286 | 0.723 | 0.078 |
| Week 12 | 2.411 | 1.173 | 0.042 | 1.091 | 0.855 | 0.204 | -1.929 | 0.739 | 0.010 |
| Age | -0.862 | 0.382 | 0.028 | 0.056 | 0.288 | 0.847 | 0.222 | 0.334 | 0.509 |
| Years since IBS diagnosis | 0.099 | 0.288 | 0.731 | 0.260 | 0.215 | 0.230 | -0.042 | 0.234 | 0.857 |
| Female | -1.210 | 2.248 | 0.592 | -0.460 | 1.685 | 0.786 | 0.858 | 1.851 | 0.644 |
|  | Estimate | SE | p | Estimate | SE | p | Estimate | SE | p |
| White | -3.641 | 2.464 | 0.145 | 3.117 | 1.856 | 0.099 | -2.551 | 2.158 | 0.242 |
| Not Hispanic/Latino | -2.286 | 3.307 | 0.492 | 1.145 | 2.492 | 0.648 | -2.225 | 2.913 | 0.448 |
| Student | 1.555 | 1.540 | 0.314 | -0.455 | 1.134 | 0.689 | 2.307 | 1.057 | 0.030 |
| clusterB:factor(visit)6 | -8.449 | 1.742 | <0.001 | 0.755 | 1.269 | 0.553 | 4.152 | 1.096 | <0.001 |
| clusterB:factor(visit)12 | -7.578 | 1.824 | <0.001 | 1.732 | 1.329 | 0.195 | 6.035 | 1.150 | <0.001 |

Block D
| Sleep_TScore |  |  |  |
| --- | --- | --- | --- |
| Age | -0.967 | 0.356 | 0.009 |
| Years since IBS diagnosis | -0.047 | 0.271 | 0.862 |
| Female | 0.632 | 2.112 | 0.765 |
| White | -3.962 | 2.292 | 0.089 |
| Not Hispanic/Latino | 0.312 | 3.071 | 0.919 |
| Student | -1.572 | 1.504 | 0.298 |
| clusterB:factor(visit)6 | -5.259 | 1.747 | 0.003 |
| clusterB:factor(visit)12 | -3.567 | 1.828 | 0.053 |

Notes: Separate linear mixed-effects models were fitted for each outcome. Fixed effects included week, phenotype, their interaction, and covariates (age, years since IBS diagnosis, sex, race, Hispanic/Latino ethnicity, and student status); participant was included as a random intercept. Week 0 and Phenotype A were used as the reference categories.

**Figure 1.**
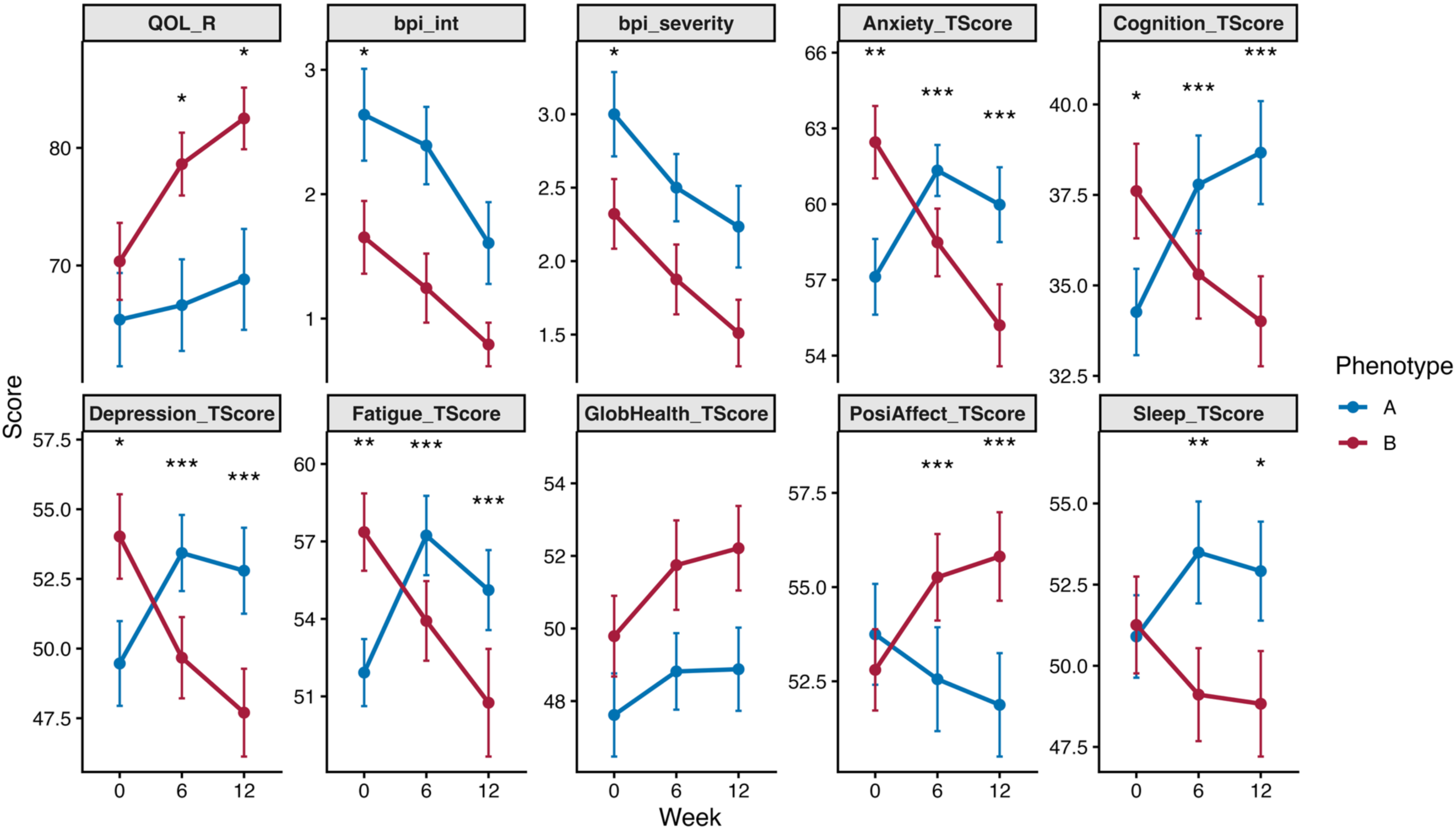
Trajectories of pain, quality of life, and psychoneurological outcomes among the two phenotypes over time. Notes: Values shown are adjusted means with standard errors, and p-values are from linear mixed-effects models controlling for sex, race, ethnicity, age, years since IBS diagnosis, and employment status, with a participant-level random intercept. At week 0, p-value annotations represent the adjusted between-phenotype difference at baseline. At weeks 6 and 12, p-value annotations represent the adjusted difference-in-differences comparing changes from baseline between phenotypes. Significance labels are defined as follows: * p < 0.05, ** p < 0.01, and *** p < 0.001. Red points and lines indicate Phenotype A, and blue points and lines indicate Phenotype B. a. IBS QOL b. BPI pain interference c. BPI pain severity d-j. Psychoneurological outcomes (anxiety, applied cognition, depression, fatigue, global health, positive affect, and sleep disturbance, and) measured by the Patient-Reported Outcomes Measurement Information System (PROMIS) Blue dots show Phenotype A, Red dots show Phenotype B.

### 3.2 Biological differences in gut microbiota composition and function among distinct IBS phenotypes

The unadjusted comparisons showed no statistically significant differences in alpha-diversity trajectories within any phenotypes (Figure 2a and Table 4). After fitting a linear mixed-effects model (LMM) adjusting for sex, race, ethnicity, age, years since IBS diagnosis, and employment status, Inverse Simpson diversity in Phenotype A showed a significant decrease at week 12 (β = −1.546, p = 0.039). No statistically significant week-12 changes were observed for Shannon or Chao1.

**Figure 2.**
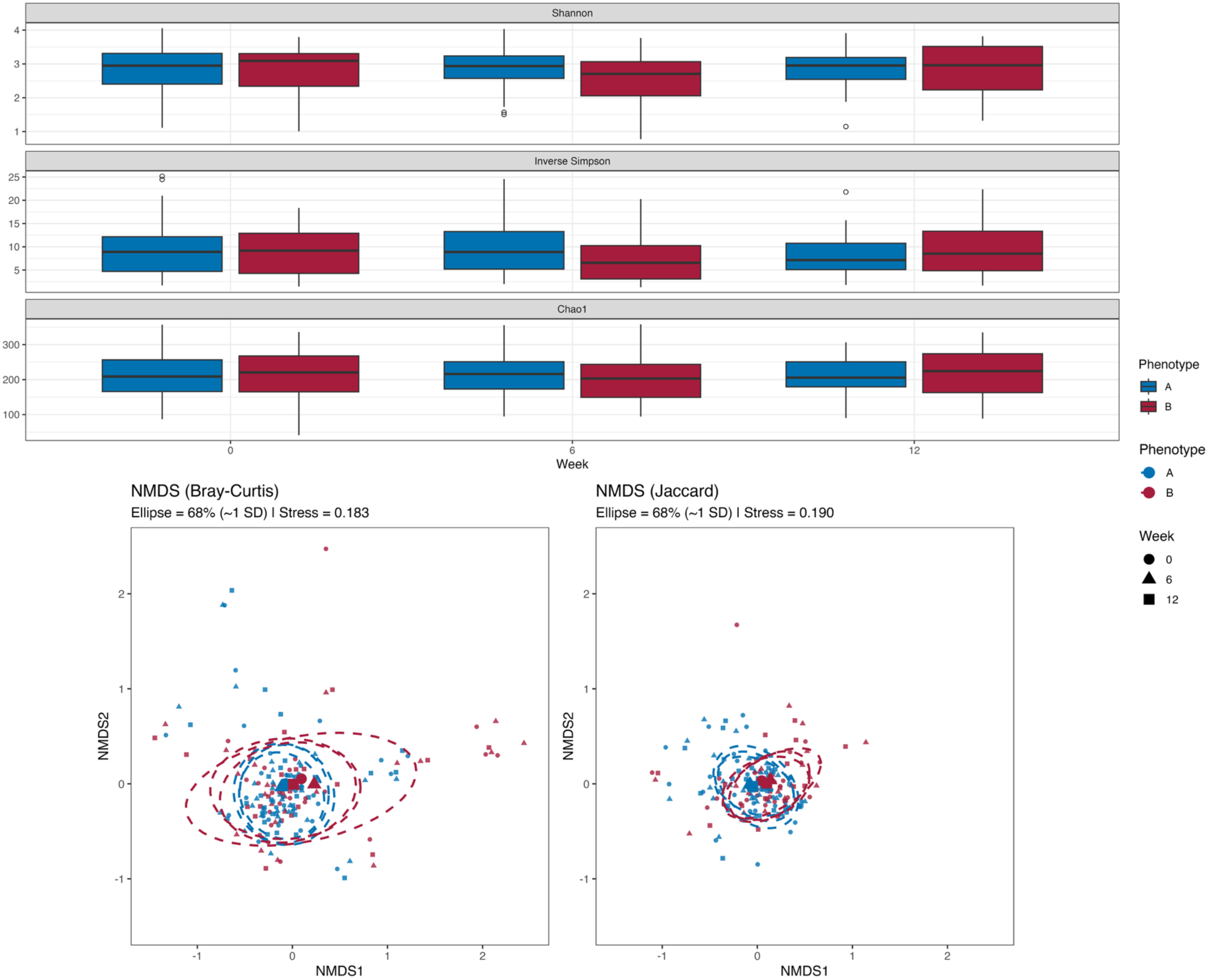
Diversity of the bacterial community among the two phenotypes over time. a.a-diversity trajectories among the two phenotypes over time. After fitting a linear mixed-effects model (LMM) adjusting for sex, race, ethnicity, age, years since IBS diagnosis, and employment status, Inverse Simpson diversity in Phenotype A showed a significant decrease at week 12 (β = −1.546, p = 0.039). No statistically significant week-12 changes were observed for Shannon or Chao1. a1. Shannon index a2. Inverse Simpson index a3. Chao index b. β-diversity trajectories among the two phenotypes over time. By PERMANOVA (adonis2; 999 permutations; marginal tests), Bray–Curtis beta diversity was not significantly associated with phenotype at baseline (R² = 0.023, p = 0.110), but was significantly associated with phenotype at week 6 (R² = 0.044, p = 0.007) and week 12 (R² = 0.030, p = 0.028), after adjusting for sex, race, ethnicity, employment status, age, and years since IBS diagnosis. Across timepoints, race was consistently associated with community composition (baseline p = 0.001; week 6 p = 0.006; week 12 p = 0.001), and years since IBS diagnosis was also associated at baseline (p = 0.032) and week 12 (p = 0.008). b1. Bray-Curtis dissimilarity; b2. Jaccard dissimilarity.

**Table 4.**
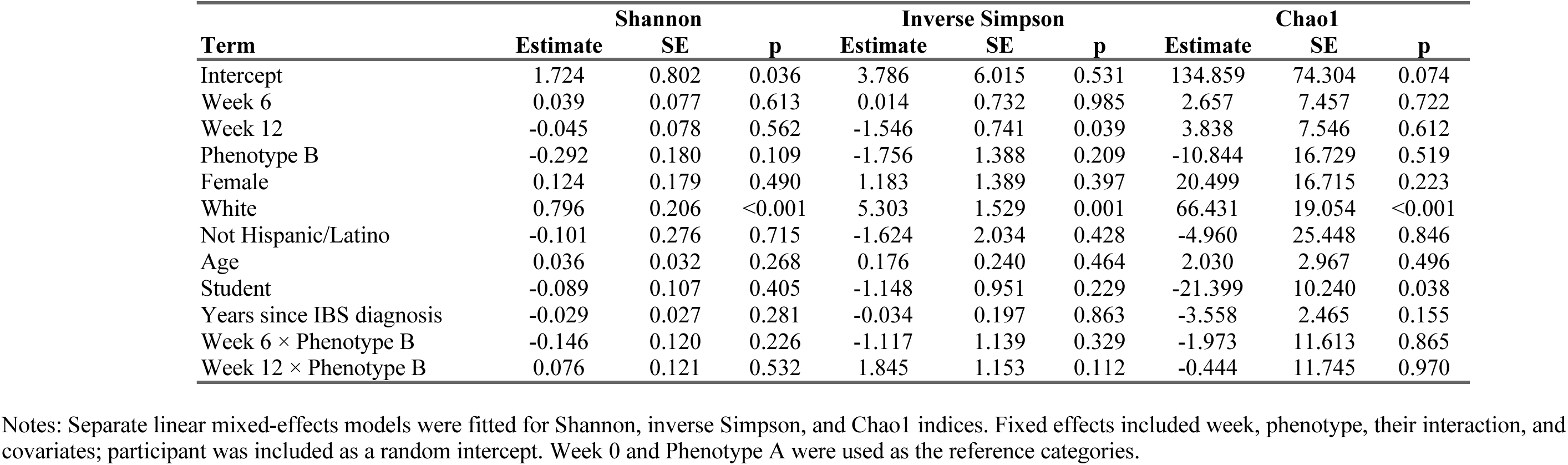
Linear mixed-effects model results for alpha-diversity indices.

In a repeated-measures PERMANOVA with permutations constrained within participant, the visit-by-phenotype interaction was also not significant (Figure 2b and Table 5). Bray–Curtis beta diversity was not significantly associated with phenotypes at baseline (R² = 0.023, p = 0.110), but was significantly associated with cluster at week 6 (R² = 0.044, p = 0.007) and week 12 (R² = 0.030, p = 0.028), after adjusting for sex, race, ethnicity, employment status, age, and years since IBS diagnosis. Across timepoints, race was consistently associated with community composition (baseline p = 0.001; week 6 p = 0.006; week 12 p = 0.001), and years since IBS diagnosis was also associated at baseline (p = 0.032) and week 12 (p = 0.008).

**Table 5.** PERMANOVA results for beta diversity by distance metric.

| Measure | Predictor | Week 0 |  | Week 6 |  | Week 12 |  | Repeated |  |
| --- | --- | --- | --- | --- | --- | --- | --- | --- | --- |
|  |  | Marginal R<sup>2</sup> | p | Marginal R<sup>2</sup> | p | Marginal R<sup>2</sup> | p | Marginal R<sup>2</sup> | p |
| **Bray-Curtis** | Phenotype | 0.023 | 0.110 | 0.044 | 0.007 | 0.030 | 0.028 |  |  |
|  | Female | 0.019 | 0.231 | 0.019 | 0.330 | 0.019 | 0.306 | 0.016 | 0.511 |
|  | White | 0.074 | 0.001 | 0.043 | 0.006 | 0.068 | 0.001 | 0.058 | 0.928 |
|  | Not Hispanic/Latino | 0.011 | 0.780 | 0.014 | 0.672 | 0.021 | 0.219 | 0.012 | 0.124 |
|  | Student | 0.017 | 0.324 | 0.016 | 0.482 | 0.015 | 0.554 | 0.009 | 0.275 |
|  | Age | 0.008 | 0.955 | 0.010 | 0.887 | 0.019 | 0.336 | 0.008 | 0.363 |
|  | Years since IBS diagnosis | 0.029 | 0.032 | 0.026 | 0.112 | 0.040 | 0.008 | 0.029 | 0.397 |
|  | Week × Phenotype |  |  |  |  |  |  | 0.005 | 0.138 |
| **Jaccard** | Phenotype | 0.020 | 0.188 | 0.034 | 0.010 | 0.028 | 0.018 |  |  |
|  | Female | 0.016 | 0.416 | 0.017 | 0.477 | 0.018 | 0.385 | 0.012 | 0.532 |
|  | White | 0.055 | 0.001 | 0.034 | 0.007 | 0.052 | 0.001 | 0.042 | 0.923 |
|  | Not Hispanic/Latino | 0.012 | 0.835 | 0.015 | 0.699 | 0.020 | 0.258 | 0.011 | 0.231 |
|  | Student | 0.016 | 0.419 | 0.016 | 0.541 | 0.017 | 0.488 | 0.009 | 0.261 |
|  | Age | 0.011 | 0.945 | 0.012 | 0.948 | 0.019 | 0.318 | 0.008 | 0.404 |
|  | Years since IBS diagnosis | 0.026 | 0.028 | 0.024 | 0.111 | 0.034 | 0.008 | 0.024 | 0.458 |
|  | Week × Phenotype |  |  |  |  |  |  | 0.006 | 0.076 |

Notes: Marginal R<sup>2</sup> and permutation p-values are shown for PERMANOVA models using 999 permutations. Cross-sectional analyses were performed separately at Week 0, Week 6, and Week 12. Repeated-measures analyses included week, phenotype, their interaction, and covariates, with permutations constrained within participant.

Exploratory analyses of longitudinal gut microbiota features among participants with available stool samples across the two phenotypes showed a compositional shift from baseline to week 12 (Figure 3). Exploratory genus-level differential abundance analyses using ANCOM-BC2 suggested between-cluster differences in trajectories (Figure 3). *Prevotella* and *Lachnobacterium* showed an estimated increase in abundance in Phenotype B, and *Megamonas* was increased in abundance in Phenotype A at baseline (Figure 3b1). At week 6 follow-up, the abundances of four genera were significantly different between the two phenotypes, with higher enrichment of *Lachnobacterium* in Phenotype A and higher enrichment of *Desulfovibrio*, *Coprobacillus*, and *Eubacterium* in Phenotype B (Figure 3b2). *Lactobacillus* showed an estimated increase in abundance in Phenotype B at week 12 (Figure 3b3). Moreover, the abundance of three genera shows compositional differences over time and across phenotypes (Figure 3c). Compared to Phenotype A, Phenotype B showed an estimated decrease in *Lachnobacterium* abundance and increased abundance of *Mitochondria-unclassified* at week 6, and a decrease in *Megasphaera* abundance at week 12.

**Figure 3.**
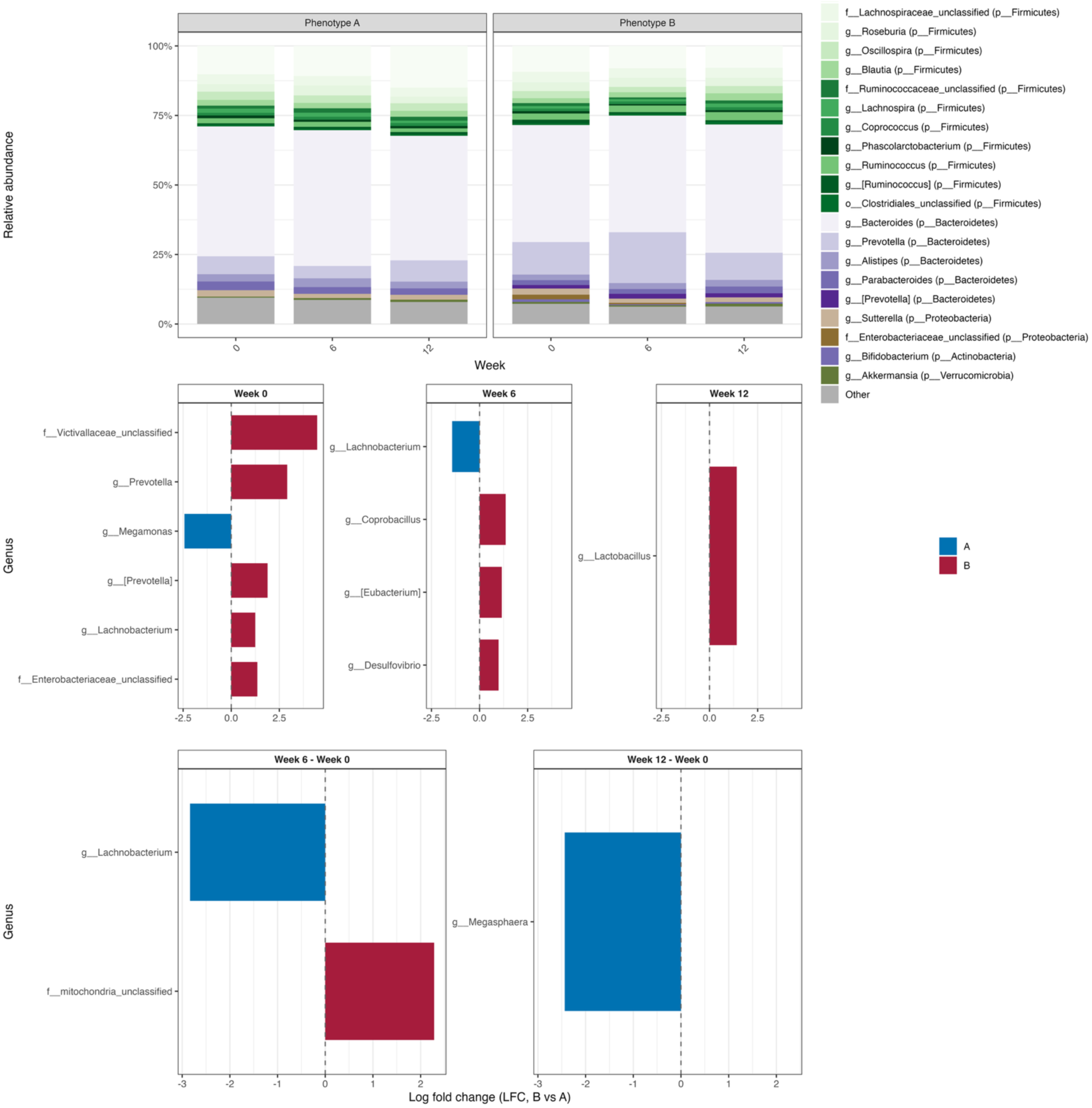
Gut microbiota compositional differences between the two phenotypes over time. (a) Relative abundance of dominant genera (colored by phylum) across Week 0, Week 6, and Week 12 for Phenotype A and Phenotype B. Stacked bar plots represent mean compositional profiles within each phenotype and time point. (b) Differential abundance analysis at each time point, shown as log fold change (LFC, Phenotype B vs. Phenotype A). Positive values indicate higher abundance in Phenotype B, while negative values indicate higher abundance in Phenotype A. The Holm procedure was used to control the false discovery rate. Only taxa with significant differences are displayed. (c) Longitudinal changes in differential abundance, represented as differences in LFC between time points (Week 6 − Week 0 and Week 12 − Week 0). These panels highlight taxa that exhibit phenotype-specific temporal shifts (i.e., interactions between phenotype and time). a. (two phenotypes over time, baseline, 6 weeks, 12 weeks) bar plot b1. difference at baseline b2. at week 6 b3. week 12 c1. Interaction effect at week 6 c2. Interaction effect at week 12

Using limma, we identified seven predicted metabolic pathways derived from gut microbiota differences in change from baseline to week 12 for KEGG functional pathways (raw p < 0.05; Figure 4), including xenobiotic degradation (ko00361/ko00623/ko00365), aromatic amino acid biosynthesis (ko00400), bile secretion (ko04976), proteasome (ko03050), and novobiocin biosynthesis (ko00401), suggesting heterogeneous functional dynamics by symptom-trajectory phenotype; although none remained significant after FDR correction. As shown in the row-scaled heatmap of the top unadjusted pathways across weeks 0, 6, and 12 (Figure 4), pathway-level changes were heterogeneous. Some pathways exhibited broadly parallel warm-to-cool shifts from baseline to week 12 in both phenotypes, while others showed transient, time-dependent fluctuations without a consistent intervention-specific separation. After adjusting for multiple testing, no pathway reached FDR significance (minimum adjusted p = 0.40); in both phenotypes, prodigiosin biosynthesis and the Caulobacter cell cycle decreased from baseline to week 12 (unadjusted p < 0.05; FDR-adjusted p > 0.05).

**Figure 4.**
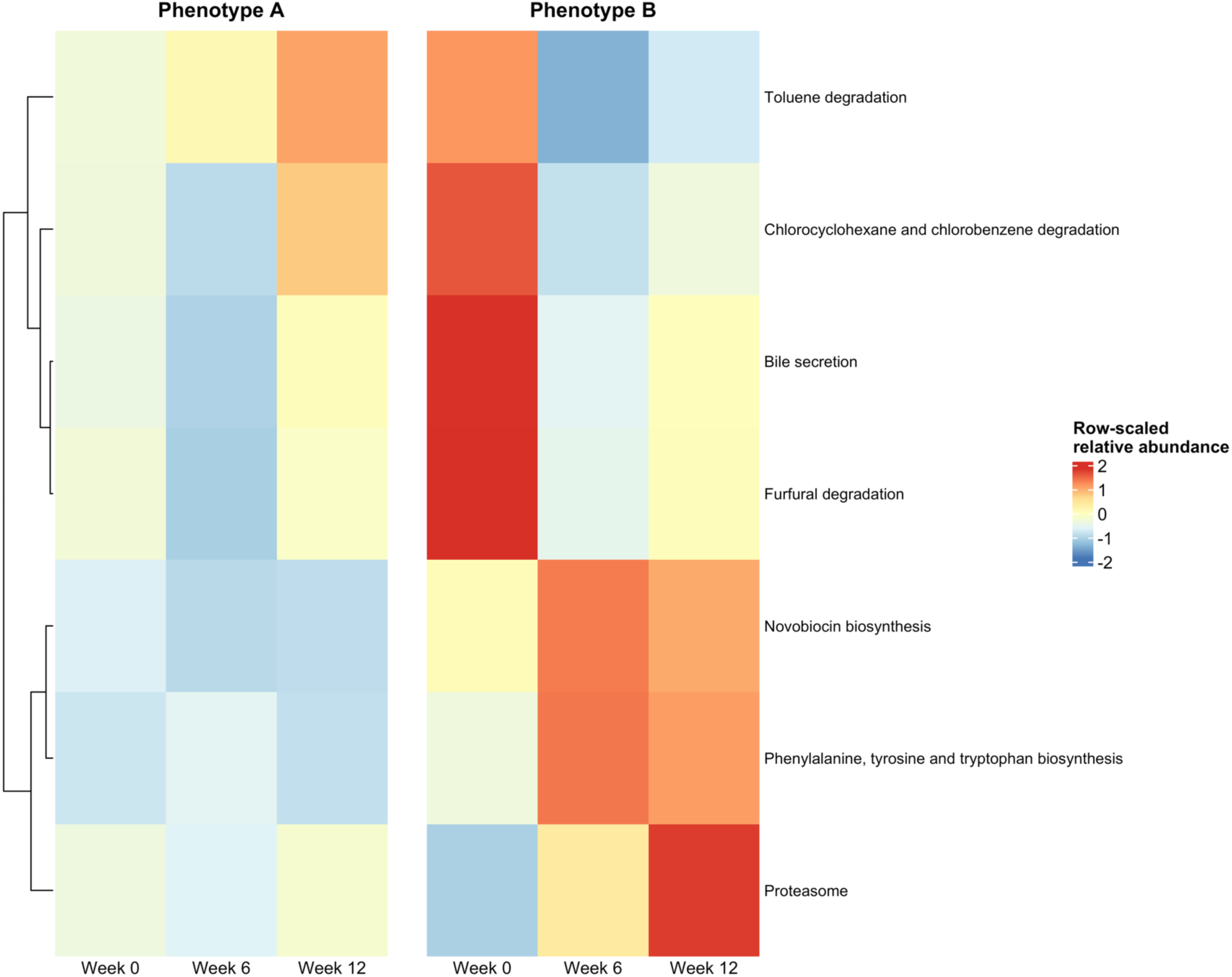
Different metabolic profiles of gut microbiota among the two phenotypes over time. Heatmap of pathway-level functional profiles across visits and phenotypes. Rows show the top pathways selected by unadjusted significance (raw p-values), and columns show the phenotype by visits combinations. Each cell represents the mean relative abundance of a pathway within that phenotype and visit, standardized within each pathway (row-wise z-score) to highlight patterns rather than absolute abundance. Warmer colors indicate time points where a pathway is above its own average level across all columns, and cooler colors indicate below its own average level. Negative values therefore reflect below-average relative abundance. Differences in KEGG pathways (level 3) were assessed using linear models with empirical Bayes moderation. The Benjamini-Hochberg procedure was used to control the false discovery rate for multiple test corrections. The p-value was set as 0.05. 7 microbial pathways differed between two clusters (raw p<0.05); none survived FDR.

### 3.3 Gut microbiota can predict the outcomes of IBS pain self-management interventions

BART models were applied to identify baseline gut microbial genera and functional pathways (KEGG Level 3) that serve as influential predictors of intervention response, defined as changes in pain severity, pain interference, and quality of life at 6 and 12 weeks. Six BART models were developed, each for Phenotype A (Figure 5 and Figure 7) and Phenotype B (Figure 6 and Figure 8), including three prediction models for change from week 0 to week 6 outcomes (pain severity, pain interference, and quality of life) and three corresponding models for week 12 outcomes. Phenotype-specific models (Phenotype A and Phenotype B) were used to characterize distinct microbiome–phenotype response profiles. To address sparsity and zero inflation in high-dimensional microbiome data, a prevalence-based filtering step was applied within each cluster, retaining features present in >10% of samples. For genus-level taxa, presence was defined as abundance >0, whereas for Tax4Fun2-predicted KEGG pathways, presence was defined using a minimum relative abundance threshold (5e-3). The RMSEs of these 12 models ranged from 8.011 to 13.965 for QOL and from 0.858 to 1.714 for pain (Table 6), indicating moderate discriminative ability. Across models, full-sample RMSEs were slightly lower than cross-validated out-of-sample RMSEs, and in-sample estimates closely approximated full-sample performance (Table 6), suggesting minimal overfitting.

**Figure 5.**
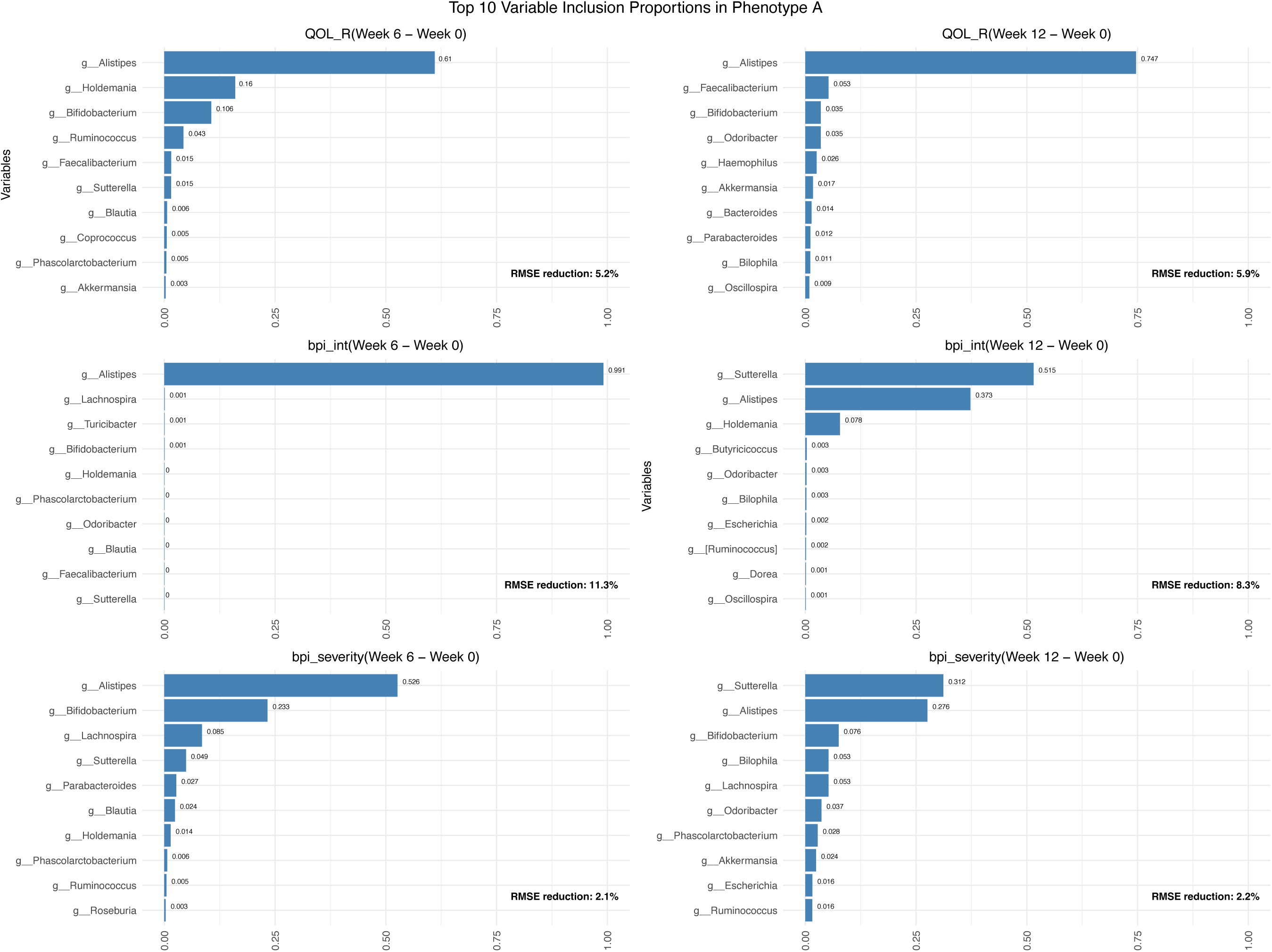
The top 10 genera of baseline gut microbiota predict intervention response, defined as changes in pain severity, pain interference, and quality of life at 6 and 12 weeks in Phenotype A.

**Figure 6.**
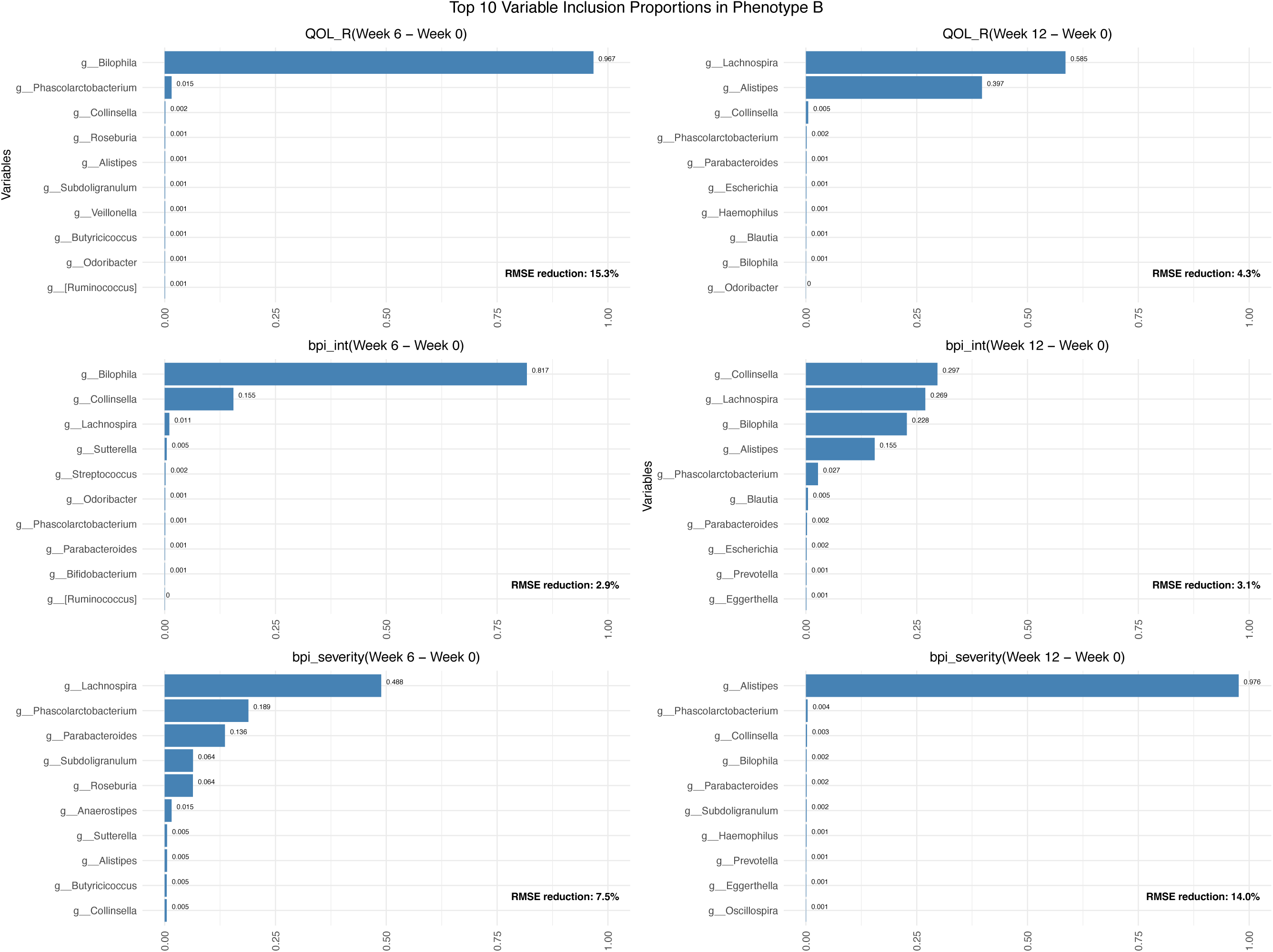
The top 10 genera of baseline gut microbiota predict intervention response, defined as changes in pain severity, pain interference, and quality of life at 6 and 12 weeks in Phenotype B.

**Figure 7.**
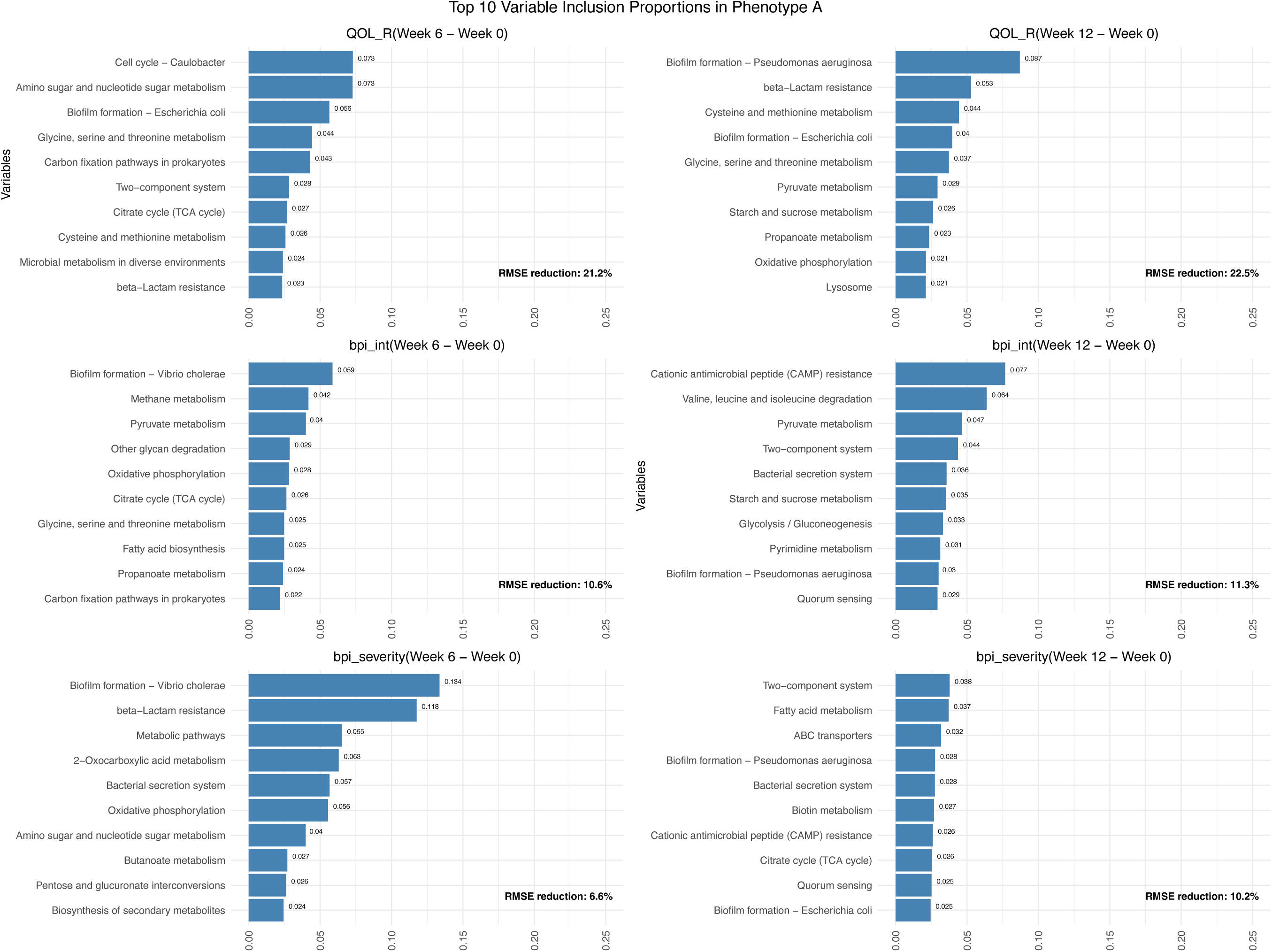
The top 10 metabolic pathways of baseline gut microbiota that predict intervention response, defined as changes in pain severity, pain interference, and quality of life at 6 and 12 weeks in Phenotype A.

**Figure 8.**
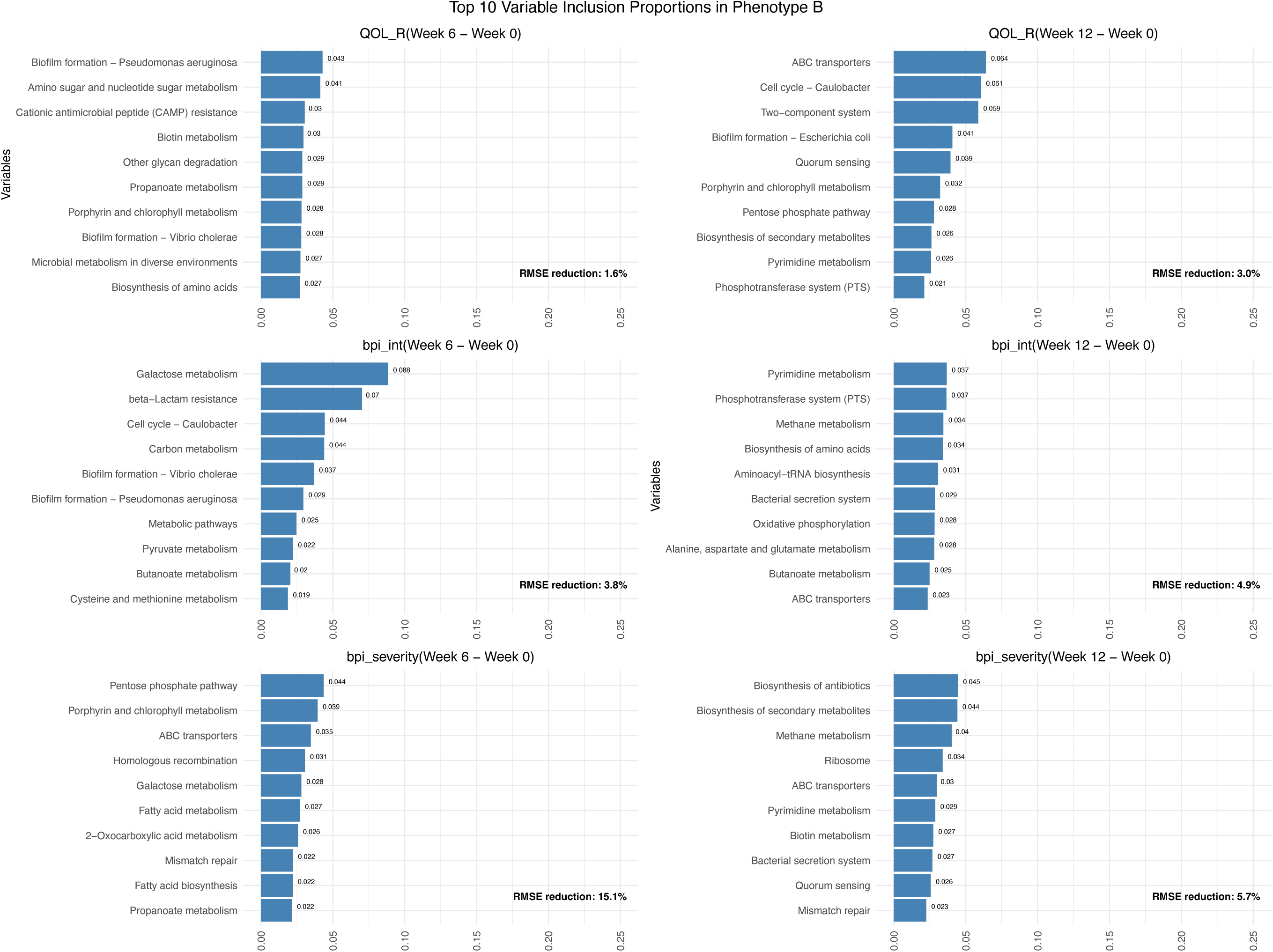
The top 10 metabolic pathways of baseline gut microbiota predict intervention response, defined as changes in pain severity, pain interference, and quality of life at 6 and 12 weeks in Phenotype B.

**Table 6.** RMSE summary by feature type and phenotype based on the BART models.

| Model | Outcome | Week 6 - Week 0 | Week 12 - Week 0 |
| --- | --- | --- | --- |
| **Genus - Phenotype A** | QOL_R | 9.212 (8.825, 10.776) | 13.965 (14.890, 15.745) |
|  | bpi_int | 1.660 (1.765, 2.153) | 1.714 (1.768, 1.951) |
|  | bpi_severity | 1.274 (1.240, 1.372) | 1.457 (1.481, 1.567) |
| **Genus - Phenotype B** | QOL_R | 8.011 (8.814, 8.687) | 9.741 (9.734, 9.982) |
|  | bpi_int | 1.164 (1.167, 1.244) | 1.168 (1.185, 1.296) |
|  | bpi_severity | 0.858 (0.898, 0.915) | 1.025 (1.098, 1.276) |
| **Pathway - Phenotype A** | QOL_R | 7.655 (8.784, 9.769) | 11.503 (12.868, 14.880) |
|  | bpi_int | 1.675 (1.760, 2.005) | 1.658 (1.685, 1.829) |
|  | bpi_severity | 1.215 (1.247, 1.374) | 1.338 (1.349, 1.696) |
| **Pathway - Phenotype B** | QOL_R | 9.306 (9.401, 10.104) | 9.870 (10.112, 10.351) |
|  | bpi_int | 1.154 (1.136, 1.229) | 1.147 (1.115, 1.317) |
|  | bpi_severity | 0.787 (0.846, 0.913) | 1.123 (1.125, 1.411) |

Notes: 3-fold cross-validation was used to select the best hyperparameter configuration for each outcome-task combination (row) based on mean out-of-sample RMSE across visits (columns). The selected configuration was then applied to the full dataset to obtain the final RMSE. Each cell shows the full-data RMSE, with mean in-sample and out-of-sample RMSEs in parentheses.

The top 10 baseline gut microbial genera identified by variable importance measures varied across models and phenotypes (Figures 5 and 6). In Phenotype A, *Alistipes* and *Sutterella* were consistently present across all models, indicating robust and reproducible microbiome predictors of intervention response; *Bifidobacterium* and *Ruminoccoccus* were also included in most models (Figure 5). In Phenotype B, *Phascolarctobacterium* was consistently present in all models, followed by *Alistipes, Collinsella*, and *Parabacteroides,* each present in 5 models (Figure 6).

The top 10 baseline gut microbiota pathways that predicted intervention response also differed across phenotypes and visits (Figures 7 and 8). In Phenotype A, the pathways “Glycine, serine and threonine metabolism” and “Cysteine and methionine metabolism” were consistently among the predictors of changes in QOL from baseline to follow-ups; “Pyruvate metabolism” consistently predicted changes in BPI pain interference; and “Bacterial secretion system” consistently predicted changes in BPI severity (Figure 7). Similar but distinct patterns in Phenotype B: only “Porphyrin and chlorophyll metabolism” was consistently among the predictors of changes in QOL from baseline to follow-ups; “Butanoate metabolism” consistently predicted changes in BPI pain interference; “Mismatch repair” consistently predicted changes in BPI severity (Figure 8).

## 4. Discussion

This analysis identified phenotype-specific mechanisms of the gut microbiome involved in pain self-management programs in young adults with IBS. Although both interventions were effective in improving patient-centered outcomes,^26^ patients did not respond uniformly; instead, they segregated into distinct multidimensional response phenotypes that were partially independent of treatment assignment. These findings indicate that intervention effects unfold within heterogeneous regulatory contexts, characterized by distinct psychoneurological and microbiome-linked response trajectories. While primary analyses demonstrated overall intervention efficacy, clustering revealed substantial heterogeneity, identifying phenotypes that may reflect underlying differences in regulatory capacity and gut–brain axis function.^15,16,42^

Building on this, our findings show that gut microbiota not only contribute to variability in outcomes but also display phenotype-specific predictive signatures. Baseline microbial genera were used to identify distinct microbiome-phenotype response profiles, suggesting that the microbial community drives variation in responses to interventions and may be used to inform optimal interventions at the individual level. In Phenotype A, genera such as *Alistipes* and *Sutterella* were consistently identified across all six models (Figure 5), with *Bifidobacterium, Lachnospira*, and *Ruminococcus* also frequently contributing to the changes in BPI severity, indicating a relatively stable and reproducible microbial signature. Increased *Alistipes* has been identified as pathogenic in colorectal cancer and is linked to the risk of depression,^43^ while *Bifidobacterium* is thought to offer health benefits.^44^ Previous studies have confirmed that higher levels of *Ruminococcus* and *Sutterella* are associated with increased IBS pain and a greater risk of cognitive decline.^45–47^ These abundance features align with the symptom trajectory in Phenotype A, which shows improvements in pain but worsening in several psychoneurological domains at 6 and 12 weeks, such as depression and applied cognition (Figure 1).

In contrast, Phenotype B was characterized by a different set of predictors, with *Phascolarctobacterium* consistently present and *Alistipes*, *Collinsella*, and *Parabacteroides* appearing in most models (Figure 6). Although the RMSE of *Phascolarctobacterium, Collinsella,* and *Parabacteroides* in the BART models was small, previous studies have confirmed their health benefits in IBS, including reducing inflammation, improving intestinal barrier integrity, and modulating the microbiota-gut-brain axis.^48–51^ This divergence suggests that different microbial arrangements may lead to similar clinical improvements, indicating multiple biological pathways that result in comparable symptom outcomes. The phenotype-response may guide precision health strategies for IBS self-management by targeting gut microbiomes underlying the condition.

A similar pattern was observed at the functional level, where predicted microbial metabolic pathways differed across phenotypes and outcomes. In Phenotype A, pathways related to amino acid metabolism (e.g., glycine, serine, threonine; cysteine and methionine) were consistently associated with improvements in quality of life, while pyruvate metabolism and bacterial secretion systems were linked to changes in pain interference and severity, respectively (Figure 7). These functional pathways regulate tryptophan metabolism, an essential amino acid, thereby affecting host physiology and immunity, linking microbial ecology with neuroimmune signaling and gut–brain interactions.^52,53^ In contrast, Phenotype B demonstrated a distinct functional profile, with pathways such as porphyrin metabolism, butanoate metabolism, and mismatch repair emerging as consistent predictors of quality of life and pain outcomes (Figure 8). These findings suggest that functional capacity of the microbiome, rather than taxonomic composition alone, may differentially shape symptom trajectories across phenotypes, potentially through effects on host metabolic, immune, and neuroregulatory processes.

Together, these results extend prior work by demonstrating that microbiome-associated mechanisms of treatment response are not uniform but instead operate through phenotype-specific microbial and functional pathways. While pathway-level differences did not remain statistically significant after multiple testing correction, their consistency across models, alignment with known IBS-related biological processes, and convergence at both taxonomic and functional levels support their biological plausibility.^54–56^ These findings highlight the importance of integrating machine learning–derived feature selection with theory-driven interpretation to uncover meaningful biological signals in high-dimensional microbiome data.

The predictive performance of machine learning models provides further support for the role of the gut microbiome in regulating IBS symptoms. Baseline microbial features were informative in predicting both symptom severity and longitudinal changes, suggesting that microbiome profiles may serve as candidate biomarkers for stratifying patients and tailoring self-management strategies. Importantly, the variability in selected features across phenotypes underscores the need for precision approaches that account for underlying heterogeneity rather than assuming a single uniform microbiome signature of response.

These findings align with emerging multi-omics evidence demonstrating coordinated alterations in gut microbial function and host biological systems in IBS.^57–59^ Within this framework, self-management interventions may act as modulators of a broader regulatory network linking microbial metabolism, host immune activity, and neuroendocrine function. The observed phenotype-specific microbiome signatures further suggest that intervention effects may be mediated through different regulatory pathways across individuals, consistent with a systems-based model of gut–brain axis dysregulation.

### 4.1 Limitations and Future Directions

Several limitations should be considered. First, functional profiles were inferred from 16S rRNA gene data rather than directly measured using shotgun metagenomics, which limits resolution and may introduce bias in predictions. Second, the sample size was modest, reducing statistical power to detect pathway-level differences after multiple-comparison correction and limiting generalizability. Third, IBS subtype classification was not incorporated, which may contribute to additional heterogeneity in symptom patterns and microbiome associations.

The variability in identified microbial features across models highlights the complexity and individualized nature of host–microbiome interactions in IBS. Future studies should focus on identifying reproducible functional pathways and integrating multi-omics approaches, including metagenomics and host transcriptomics, to better characterize mechanistic links. Larger, longitudinal cohorts are needed to validate trajectory phenotypes and microbial predictors, as well as to examine within-person dynamics over time.

From a translational perspective, these findings support the development of stratified care models in IBS. Future research should evaluate whether tailoring interventions based on microbiome and symptom profiles improves clinical outcomes. Mechanism-informed approaches targeting specific regulatory pathways—such as behavioral interventions, neuromodulation, or pharmacologic strategies—may enhance treatment precision and effectiveness.

## 5. Conclusion

In summary, this study identified distinct symptom trajectory phenotypes in young adults with IBS and demonstrated that gut microbiota composition and functional profiles are associated with differential responses to a pain self-management intervention. These findings highlight the potential of microbiome-informed stratification to advance personalized, mechanism-based approaches for IBS management.

## Author Contributions

Conceptualization, J.C., M.H.C., and X.C.; formal analysis, J.C., A.L., W.W., Z.T., M.H.C., and X.C.; funding acquisition, X.C., and A.S.; methodology, J.C., A.L., W.W., Z.T., W.X., A.S., M.H.C., and X.C.; project administration, J.C., W.X., A.S., and X.C.; writing-original draft, J.C., A.L., and X.C.; writing-review and editing, All. All authors have reviewed the manuscript and agreed to submit this version.

## Acknowledgments

The authors would like to acknowledge all the participants in this study. The authors would also like to acknowledge the support from the Bio-Behavioral Lab (BBL), the Center of Advancement in Managing Pain (CAMP), and the NIH-funded P20 Center for Accelerating Precision Pain Self-Management in the University of Connecticut School of Nursing. The authors would like to acknowledge the support from the Microbial Analysis, Resources, and Services (MARS) and the Institute for Systems Genomics (ISG) at the University of Connecticut.

## Data Availability Statement

The datasets generated and analyzed in the current study are available from the NCBI SRA (https://submit.ncbi.nlm.nih.gov/subs/sra/SUB8914789/). Deidentified data will be available upon reasonable request. Requests to access these datasets should be directed to.

